# Germany’s low SARS-CoV-2 seroprevalence confirms effective containment in 2020: Results of the nationwide RKI-SOEP study

**DOI:** 10.1101/2021.11.22.21266711

**Authors:** Hannelore Neuhauser, Angelika Schaffrath Rosario, Hans Butschalowsky, Sebastian Haller, Jens Hoebel, Janine Michel, Andreas Nitsche, Christina Poethko-Müller, Franziska Prütz, Martin Schlaud, Hans W. Steinhauer, Hendrik Wilking, Lothar H. Wieler, Lars Schaade, Stefan Liebig, Antje Gößwald, Markus M. Grabka, Sabine Zinn, Thomas Ziese

## Abstract

Pre-vaccine SARS-CoV-2 seroprevalence data from Germany are scarce outside hotspots, and socioeconomic disparities remained largely unexplored. The nationwide RKI-SOEP study with 15,122 adult participants investigated seroprevalence and testing in a supplementary wave of the Socio-Economic-Panel conducted predominantly in October-November 2020. Self-collected oral-nasal swabs were PCR-positive in 0.4% and Euroimmun anti-SARS-CoV-2-S1-IgG ELISA from dry capillary blood in 1.3% (95% CI 0.9-1.7%, population-weighted, corrected for sensitivity=0.811, specificity=0.997). Seroprevalence was 1.7% (95% CI 1.2-2.3%) when additionally adjusting for antibody decay. Overall infection prevalence including self-reports was 2.1%. We estimate 45% (95% CI 21-60%) undetected cases and analyses suggest lower detection in socioeconomically deprived districts. Prior SARS-CoV-2 testing was reported by 18% from the lower educational group compared to 25% and 26% from the medium and high educational group (p<0.0001). Symptom-triggered test frequency was similar across educational groups. However, routine testing was more common in low-educated adults, whereas travel-related testing and testing after contact with an infected person was more common in highly educated groups. In conclusion, pre-vaccine SARS-CoV-2-seroprevalence in Germany was very low. Notified cases appear to capture more than half of infections but may underestimate infections in lower socioeconomic groups. These data confirm the successful containment strategy of Germany until winter 2020.

## Introduction

The first case of severe acute respiratory syndrome coronavirus 2 (SARS-CoV-2) infection in Germany was reported on January 27^th^ 2020. In line with the National Pandemic Response Plan, early on in 2020 high testing capacities, a containment strategy with regulations on physical distancing and movement as well as closures of daycare facilities, schools and in the retail sector were established^1^. In spring 2020, case counts in Germany were relatively low, but the proportion of the population infected was supposed to be much higher due to asymptomatic or mild symptomatic cases that might not have been captured by the predominantly symptom-triggered testing. One of the first highly publicized seroepidemiologic studies worldwide was carried out in the small German hotspot of Gangelt ^2^. This study reported that 16% of the Gangelt population had been infected by early April 2020 and concluded that undetected cases account for about 80% of total cases. Several reports of a SARS-CoV-2 seroprevalence around 10% in the spring and summer 2020 from other hotspot areas in Germany followed ^3,4^. Although far from representing the German population, these initial reports of high seroprevalence in local hotspots raised the expectation of an already high, albeit unknown, countrywide proportion of the population with antibodies against SARS-CoV-2. In contrast, a still low seroprevalence outside hotspots was suggested by testing blood donors^5,6^ and by a population-based cohort from the city of Bonn, which all showed a SARS-CoV-2 IgG seroprevalence below 1% by June 2020^7^. In line with this, a study conducted in Munich, in the more highly affected south of Germany, found that seroprevalence by the beginning of summer was at only 1.8%^8^.

To obtain national estimates of cumulative SARS-CoV-2 infections in Germany in the late fall of 2020, the Robert Koch Institute (RKI) initiated a large nationwide study as part of its seroepidemiological studies program CORONA MONITORING. In recognition of the importance of the socio-economic determinants and consequences of the pandemic, the nationwide RKI-SOEP study was based on the representative dynamic cohort German Socio-Economic Panel (SOEP). The SOEP offers comprehensive longitudinal data on the sociodemographic background and the living conditions of its participants. We present data on the prevalence and determinants of anti-SARS-CoV-2-IgG seropositivity among adults in Germany before the beginning of the vaccination roll-out. Furthermore, self-reports of SARS-CoV-2-testing and reasons for testing are analyzed in order to assess test strategies and surveillance data of the pandemic in 2020.

## Methods

### Study design and study population

The study design, its population and recruitment of participants are described in detail in the study protocol ^9^. In brief, the study methods were guided by the WHO protocol for population-based age-stratified seroepidemiological investigations for coronavirus 2019 (COVID-19) infection ^10^. The study was designed as an extraordinary wave of a dynamic population-based cohort study, the Socio-Economic Panel (SOEP). The SOEP is a nationwide longitudinal multidisciplinary household survey based on an established multistage random sampling design. In this study, since 1984 persons in private households have been asked annually about a variety of topics. These include socio-demographic characteristics, income, labor market participation and family situation, health, and their basic orientations, concerns and satisfactions. Adults from the entire SOEP gross sample in 2020, i.e. 19,569 households with N=31,675 adults, were invited to participate in the RKI-SOEP study. This gross sample covers all 401 districts in Germany. Approval was obtained from the ethics committee of the Berlin Doctors’ Council (reference ID Eth-33/20). The RKI-SOEP study was carried out in accordance with the Declaration of Helsinki. All participants provided informed consent.

### Data collection and laboratory methods

Invitations and study materials were sent by mail and included a one-page questionnaire, a self-sampling kit for dry capillary blood (DBS), a self-sampling kit for an oral-nasal swab (ONS) sample for PCR testing, illustrated instructions and a link to video material on self-sampling and FAQs^9^. Study participants were asked to take several dried blood spots (DBS) by capillary finger prick and an ONS ^9^. Both were sent by mail to the RKI. From ONS, RNA was extracted with the QIAamp Viral RNA Mini Kit (Qiagen, Hilden, Germany) and tested by real-time reverse transcription polymerase chain reaction (PCR), targeting the E gene and the orf1ab region of SARS-CoV-2^11^. The PCR was regarded positive when both targets tested positive. Standardized punches of DBS (DBS Puncher, PerkinElmer, Waltham MA, USA) were extracted according to the manufacturer’s protocol (Euroimmun AG, Lübeck, Germany) and tested for SARS-CoV-2 anti-S1 IgG antibodies using lots E200518BC (from Oct 12th to Dec 2nd, 2020) and E200831BC (from Dec 3rd, 2020 to end of study) of the Anti-SARS-CoV-2-ELISA (IgG) (Euroimmun AG, Lübeck, Germany). For defining seropositivity, the ratio cutpoint provided by the manufacturer was adapted from 1.1 to 0.94 for DBS testing (see **Supplement**). Indeterminate results were considered negative. All analyses were done on a EUROLabWorkstation ELISA (Euroimmun AG, Lübeck, Germany), testing 5 quality control specimens (3 provided by the manufacturer, 2 pooled serum controls with ratios of 1-2 and 2-3, respectively) on each 96-well plate. All 3 round robin tests on SARS-CoV-2 IgG antibodies of the INSTAND interlaboratory comparison program (INSTAND, Düsseldorf, Germany) were passed. The two RKI laboratories involved in ONS and DBS analyses are accredited laboratories according to DIN EN ISO 17025 and/or DIN EN ISO 15189 (Deutsche Akkreditierungsstelle, Frankfurt/Main, Germany). Both RKI laboratories have successfully participated in external quality assessments (EQAs) on the detection of SARS-CoV-2 genome and/or SARS-CoV-2 IgG antibodies, offered by INSTAND (INSTAND, Düsseldorf, Germany).

Self-reported questionnaire information on pre-study SARS-CoV-2 testing refers to tests based on nasal or oral swabs, excluding self-tests. Participants reporting a positive test are described as “aware of infection” in this paper, participants reporting a negative or no test are “unaware of infection”. The overall infection status was considered positive if at least one of the three indicators (PCR result from the ONS, IgG antibody result from the DBS, or self-reported pre-study SARS-CoV-2 test) was positive. It was considered negative when all available results were negative. At least one of the three indicators had to be available; sensitivity analyses requiring complete data yielded similar results.

Household composition was classified for participants aged < 60 years based on the total number of household members and whether or not there were children ≤ 16 years living in the household. To avoid confounding by age, this variable was not defined for participants ≥ 60 years, since there are practically no households with children in this age group within the study population. School education was classified as low (school dropout or low secondary school graduation), middle (intermediate secondary school graduation) or high (university entrance qualification), based on data collected in previous SOEP waves. The place of living was classified at the district level, firstly, using an index of socioeconomic deprivation with three categories (low: quintile 1, medium: quintiles 2–4, high: quintile 5)^12^, and secondly, classified into four incidence strata (high incidence, average incidence, late second wave, low incidence) based on the temporal pattern of notified SARS-CoV-2 cases in the district, as described in the **Supplement**.

### Statistical analysis

Unless stated otherwise, all analyses are weighted to allow generalizing the findings to the general adult population and to counteract non-response bias. For example, smokers and persons with non-German nationality participated significantly less often than non-smokers and persons with a German nationality. The weighting factors result from complex modelling of contactability and participation probabilities. A total of about 400 characteristics at the person and household level (taken from previous waves of the SOEP) were reviewed for inclusion in the different weighting steps. The characteristics reviewed include socio-demographic characteristics, characteristics on health status, housing situation and attitudes, e.g. towards political measures regarding COVID-19 containment. Furthermore, the weights were calibrated to national statistics at the person level (age and gender distribution of adult persons in private households) and at the household level (number of households by federal state, municipality size, household size and home ownership). The sampling and weighting have been described in detail^13^.

IgG seroprevalence was corrected for the test characteristics, i.e. sensitivity and specificity ^14^. The sensitivity (81.1%, 95% CI 77.3 - 84.4%) and specificity (99.7%, 95% CI 98.7 - 99.9%) of the Euroimmun test were estimated in validation studies by the Paul Ehrlich Institute (personal communication, H. Scheiblauer, 16 Mar 2021). For specificity, these studies comprised 576 pre-pandemic plasma samples from Germany and 100 pre-pandemic serum samples from US blood donors. For initial test sensitivity, three seroconversion cohorts were combined including a COVID-19-patient sample from University Hospital Frankfurt (n = 271), 100 patients from a commercially available seroconversion panel and 100 US patients. As the generalizability of such estimates to the given study population depends on factors such as sex, age, severity of infection and time since infection, in addition to these initial test characteristics we also corrected for sensitivity as estimated within the study, i.e. the probability of seropositivity among study participants with a self-reported positive SARS-CoV-2 test at least 11 days pre-study.

The number of infections missed by the mandatory notification system was estimated in two ways: first internally, by estimating the proportion of seropositive cases that was unaware of the infection (**Table 2**) and second by comparing the seroprevalence observed in the study, adjusted for test characteristics, to the number of notified cases, adjusted for sampling density (**Table 3 and Table S3**). Methodological details and sensitivity analyses based on three different assumptions on antibody decay are presented in the **Supplement**.

**Table 1:** Characteristics and SARS-CoV-2 infection status in community-dwelling adults in Germany (15,122 RKI-SOEP study participants, predominantly October-November 2020)

|  | Total | PCR+ self-reported | PCR+ during study | Sero-positive | SARS-CoV-2 infections |  |  |
| --- | --- | --- | --- | --- | --- | --- | --- |
|  |  |  |  |  | All infections |  |  |
|  | N | N | N | N | N (row %, unweighted) | Row % (95% CI), population-weighted | Odds Ratio and p-value, adjusted* (95% CI) |
| **Total 18-99 years** | 15,121 | 146 | 51 | 192 | 288 (1.9%) | 2.1% (1.6 - 2.6) |  |
| **Total 18-69 years** | 12,582 | 137 | 41 | 167 | 252 (2.0%) | 2.3% (1.8 - 2.9) |  |
| **Sex** |  |  |  |  |  |  | *p* = 0.59 |
| **Women** | 8,099 | 69 | 28 | 82 | 138 (1.7%) | 2.2% (1.6 - 2.8) | Ref. |
| **Men** | 7,022 | 77 | 23 | 110 | 150 (2.1%) | 2.0% (1.5 - 2.6) | 0.91 (0.64 - 1.29) |
| **Age group** |  |  |  |  |  |  | *p* = 0.38 |
| 18-34 years | 2,804 | 38 | 8 | 46 | 65 (2.3%) | 2.8% (1.9 - 4.2) | Ref. |
| 35-49 years | 3,553 | 38 | 16 | 35 | 70 (2.0%) | 2.3% (1.6 - 3.4) | 0.71 (0.42 - 1.20) |
| 50-64 years | 4,945 | 52 | 15 | 68 | 92 (1.9%) | 2.0% (1.3 - 2.9) | 0.85 (0.48 - 1.49) |
| 65-79 years | 3,126 | 18 | 9 | 39 | 55 (1.8%) | 1.3% (0.8 - 2.0) | 0.57 (0.31 - 1.04) |
| 80-99 years | 693 | - | 3 | 4 | 6 (0.9%) | 0.9% (0.3 - 2.6) | 0.51 (0.15 - 1.71) |
| **Household composition (< 60 y)** |  |  |  |  |  |  | *p* = 0.028 |
| 1 person | 1,198 | 7 | 4 | 10 | 17 (1.4%) | 1.8% (1.0 - 3.4) | Ref. |
| 2-4 persons, incl. children ≤ 16 y | 3,572 | 36 | 16 | 38 | 66 (1.8%) | 2.2% (1.5 - 3.3) | 1.00 (0.44 - 2.29) |
| 2-4 persons, no children ≤ 16 y | 3,471 | 46 | 5 | 52 | 75 (2.2%) | 2.0% (1.3 - 3.0) | 0.92 (0.40 - 2.11) |
| > 4 persons, incl. children ≤ 16 y | 1,192 | 23 | 9 | 24 | 40 (3.4%) | 6.1% (2.9 - 13) | 3.38 (1.25 - 9.16) |
| > 4 persons, no children ≤ 16 y | 113 | 2 |  | 2 | 2 (1.8%) | 0.7% (0.2 - 3.0) | -- |
| Not classified (60+ years) |  |  |  |  |  |  |  |
| **School education** |  |  |  |  |  |  | *p* = 0.18 |
| Low | 2,748 | 16 | 12 | 32 | 49 (1.8%) | 2.0% (1.3 - 3.0) | 1.31 (0.73 - 2.35) |
| Middle | 5,178 | 43 | 15 | 64 | 95 (1.8%) | 1.9% (1.4 - 2.8) | 0.77 (0.49 - 1.21) |
| High | 6,296 | 74 | 23 | 82 | 124 (2.0%) | 2.0% (1.5 - 2.7) | Ref. |
| **District incidence stratum\*\*** |  |  |  |  |  |  | *p* = 0.034 |
| High incidence (Cluster A) | 3,032 | 33 | 13 | 47 | 68 (2.2%) | 2.6% (1.7 - 4.1) | Ref. |
| Average incidence (Cluster B) | 8,688 | 92 | 29 | 117 | 176 (2.0%) | 2.1% (1.5 - 2.8) | 1.10 (0.65 - 1.86) |
| Late second wave (Cluster C) | 1,715 | 16 | 7 | 18 | 32 (1.9%) | 2.2% (1.2 - 3.9) | 0.94 (0.46 - 1.93) |
| Low incidence (Cluster D) | 1,681 | 5 | 2 | 10 | 12 (0.7%) | 0.8% (0.3 - 1.7) | 0.26 (0.10 - 0.72) |
| **Socioeconomic deprivation (district level)** |  |  |  |  |  |  | *p* = 0.007 |
| Low deprivation | 3,991 | 43 | 21 | 65 | 95 (2.4%) | 2.6% (1.9 - 3.6) | Ref. |
| Middle deprivation | 8,981 | 80 | 22 | 103 | 151 (1.7%) | 1.7% (1.2 - 2.4) | 0.85 (0.48 - 1.50) |
| High deprivation | 2,144 | 23 | 8 | 24 | 42 (2.0%) | 2.5% (1.5 - 4.0) | 2.03 (1.03 - 4.00) |

\*Odds ratio mutually adjusted for the variables in this table (with age modelled as a natural cubic spline), date of study participation (as a linear variable) and sampling batch. \*\*District incidence strata according to pattern of weekly sequence of district SARS-CoV-2 incidence (notified cases), see Supplement.

**Table 2:** Characteristics and IgG seroprevalence in community-dwelling adults in Germany (14,781 RKI-SOEP study participants with valid dried blood spot specimens, sampled predominantly in October-November 2020)

|  | Total | N sero-positive (row %, unweighted) | Proportion of seropositives, adjusted for specificity= 0.997 and initial test sensitivity = 0.811 | Estimate of cumulative seroprevalence since the beginning of the pandemic adjusted for specificity = 0.997 and sensitivity that includes antibody decay = 0.616 | Seropositive cases unaware of infection |
| --- | --- | --- | --- | --- | --- |
|  | N | N (row %) | Row % (95% CI) |  | % of seropositives (95% CI) |
| **Total 18-99 years** | 14,781 | 192 (1.3%) | 1.3% (0.9 – 1.7) | 1.7% (1.2 – 2.3) | 48% (37 – 59) |
| **Total 18-69 years** | 12,324 | 167 (1.4%) | 1.4% (1.0 – 2.0) | 1.9% (1.3 – 2.7) | 45% (34 – 57) |
| **Sex** |  |  |  |  |  |
| Women | 7,938 | 82 (1.0%) | 1.1% (0.6 – 1.7) | 1.4% (0.8 – 2.2) | 49% (32 – 67) |
| Men | 6,843 | 110 (1.6%) | 1.5% (1.0 – 2.1) | 1.9% (1.3 – 2.8) | 46% (31 – 62) |
| **Age group** |  |  |  |  |  |
| 18-34 y | 2,741 | 46 (1.7%) | 2.0% (1.1 – 3.5) | 2.6% (1.4 – 4.7) | 43% (26 – 61) |
| 35-49 y | 3,484 | 35 (1.0%) | 1.2% (0.5 – 2.4) | 1.6% (0.7 – 3.2) | 47% (22 – 73) |
| 50-64 y | 4,846 | 68 (1.4%) | 1.3% (0.7 – 2.1) | 1.7% (0.9 – 2.8) | 44% (26 – 64) |
| 65-79 y | 3,055 | 39 (1.3%) | 0.6% (0.2 – 1.3) | 0.8% (0.3 – 1.7) | 68% (40 – 87) |
| 80-99 y | 655 | 4 (0.6%) | 0.2% (0.2 – 1.7) | 0.3% (0.2 – 2.2) | 100% |
| **Household composition (< 60 y)** |  |  |  |  |  |
| 1 person | 1,168 | 10 (0.9%) | 1.0% (0.2 – 2.9) | 1.3% (0.3 – 3.8) | 31% (6.1 – 75) |
| 2-4 persons, incl. children ≤ 16 y | 3,494 | 38 (1.1%) | 1.3% (0.6 – 2.5) | 1.7% (0.8 – 3.2) | 35% (16 – 61) |
| 2-4 persons, no children ≤ 16 y | 3,410 | 52 (1.5%) | 1.1% (0.6 – 1.9) | 1.5% (0.8 – 2.5) | 48% (28 – 68) |
| > 4 persons, incl. children ≤ 16 y | 1,168 | 24 (2.1%) | 4.4% (2.0 – 9.0) | 5.8% (2.7 – 12) | 54% (20 – 85) |
| > 4 persons, no children ≤ 16 y | 112 | 2 (1.8%) | 0.5% (0.2 – 3.2) | 0.6% (0.2 – 4.2) | 0% |
| Not classified (60+ years) | 5,078 |  |  |  |  |
| **School education** |  |  |  |  |  |
| Low | 2,662 | 32 (1.2%) | 1.5% (0.7 – 2.7) | 1.9% (0.9 – 3.5) | 52% (26 – 77) |
| Middle | 5,057 | 64 (1.3%) | 1.1% (0.6 – 1.8) | 1.4% (0.8 – 2.4) | 51% (30 – 71) |
| High | 6,177 | 82 (1.3%) | 1.1% (0.7 – 1.7) | 1.4% (0.9 – 2.2) | 45% (28 – 62) |
| **District incidence stratum\*** |  |  |  |  |  |
| High incidence (Cluster A) | 2,979 | 47 (1.6%) | 2.1% (1.0 – 3.9) | 2.7% (1.3 – 5.1) | 46% (27 – 65) |
| Average incidence (Cluster B) | 8,497 | 117 (1.4%) | 1.1% (0.7 – 1.6) | 1.4% (0.9 – 2.1) | 44% (30 – 60) |
| Late second wave (Cluster C) | 1,658 | 18 (1.1%) | 1.3% (0.4 – 3.2) | 1.7% (0.5 – 4.3) | 66% (34 – 88) |
| Low incidence (Cluster D) | 1,643 | 10 (0.6%) | 0.5% (0.0 – 1.8) | 0.7% (0.0 – 2.4) | 65% (25 – 92) |
| **Socioeconomic deprivation (district level)** |  |  |  |  |  |
| Low deprivation | 3,915 | 65 (1.7%) | 1.4% (0.9 – 2.3) | 1.9% (1.1 – 3.0) | 45% (27 – 64) |
| Middle deprivation | 8,790 | 103 (1.2%) | 1.1% (0.6 – 1.8) | 1.4% (0.8 – 2.3) | 47% (32 – 63) |
| High deprivation | 2,072 | 24 (1.2%) | 1.7% (0.7 – 3.5) | 2.3% (1.0 – 4.6) | 57% (29 – 81) |

Numbers do not add up to total due to missing values. All % are population-weighted unless otherwise specified. \*District incidence strata according to pattern of weekly sequence of district SARS-CoV-2 incidence (notified cases), see Supplement.

**Table 3:** Cumulative incidence of notified cases, underascertainment ratio and estimated proportion of undetected cases.

|  | Cumulative incidence of notified cases (non-fatal)* | Underascertainment ratio*,** | Proportion of undetected cases*,*** |
| --- | --- | --- | --- |
|  |  | Ratio (95% CI) | % (95% CI) |
| **Total 18-99 years** | 0.9% | 1.8 (1.3 – 2.5) | 45% (21 - 60) |
| **Total 18-69 years** | 1.0% | 1.9 (1.3 – 2.6) | 47% (24 - 62) |
| **Sex** |  |  |  |
| **Women** | 0.9% | 1.5 (0.9 – 2.4) | 34% (-10 - 58) |
| **Men** | 0.9% | 2.1 (1.4 – 3.1) | 54% (30 - 68) |
| **Age group** |  |  |  |
| 18-34 years | 1.3% | 2.0 (1.1 – 3.5) | 50% (6.3 - 72) |
| 35-49 years | 1.1% | 1.5 (0.6 – 2.9) | 31% (-59 - 66) |
| 50-64 years | 0.8% | 2.1 (1.2 – 3.6) | 53% (16 - 72) |
| 65-79 years | 0.4% | 1.9 (0.6 – 4.1) | 48% (-62 - 76) |
| 80-99 years | 0.6% | 0.5 (-0.4 – 3.7) | NA**** |
| **District incidence stratum** |  |  |  |
| High incidence (Cluster A) | 1.4% | 2.0 (1.0 – 3.7) | 50% (-2.8 - 73) |
| Average incidence (Cluster B) | 0.9% | 1.6 (1.0 – 2.4) | 39% (4.3 - 59) |
| Late second wave (Cluster C) | 0.7% | 2.5 (0.8 – 6.3) | 60% (-31 - 84) |
| Low incidence (Cluster D) | 0.4% | 1.8 (-0.1 – 6.6) | 44% (NA**** - 85) |
| **Socioeconomic deprivation (district level)** |  |  |  |
| Low deprivation | 1.1% | 1.7 (1.0 – 2.6) | 40% (-1.2 - 62) |
| Middle deprivation | 0.9% | 1.6 (0.9 – 2.7) | 38% (-12 - 63) |
| High deprivation | 0.5% | 4.3 (1.8 – 8.7) | 77% (46 - 89) |

\*Sampling-density adjusted, i.e. each participant contributes according to the cumulative incidence of notified cases with symptom onset (notified or imputed) corresponding to his/her DBS testing date minus 14 days.
\*\*Underascertainment ratio: Ratio of seroprevalence to cumulative incidence of notified cases. Seroprevalence population-weighted IgG seroprevalence adjusted for specificity = 0.997 and sensitivity = 0.616 that includes antibody decay observed in the study.
\*\*\*Proportion of undetected cases: Seroprevalence minus cumulative incidence of notified cases, divided by seroprevalence.
\*\*\*\*NA = estimate not available

Analyses were performed with SAS 9.4 (SAS Institute Inc., Cary, NC, USA).

To account for weighting and household clustering, we calculated robust standard errors via survey procedures to estimate 95% confidence intervals (CI) for proportions (calculated on the logit scale). p-values for proportions were obtained with Rao-Scott chi-squared tests. ORs and adjusted p-values were derived from survey logistic regression. CIs for the seroprevalence with adjustment for test characteristics were derived by transforming the unadjusted confidence limits according to the correction formula, ignoring the variability in the estimates of sensitivity and specificity. Similarly, CIs for the underascertainment ratio and the proportion of undetected cases were obtained by calculating these numbers using the confidence limits for the adjusted seroprevalence, assuming the individually matched cumulative incidence of notified cases as fixed. Results are presented for the adult population (18 years and older) as well as for the population aged 18-69 years, as this younger population is less affected by COVID-19 mortality and has only a minor percentage of the population institutionalized.

## Results (with subheadings)

**Figure 1** illustrates the chronological distribution of notified SARS-CoV-2 cases in the adult population in Germany from the start of the pandemic, as well as the sampling distribution in our study which occurred predominantly in October and November 2020.

**Figure 1.**
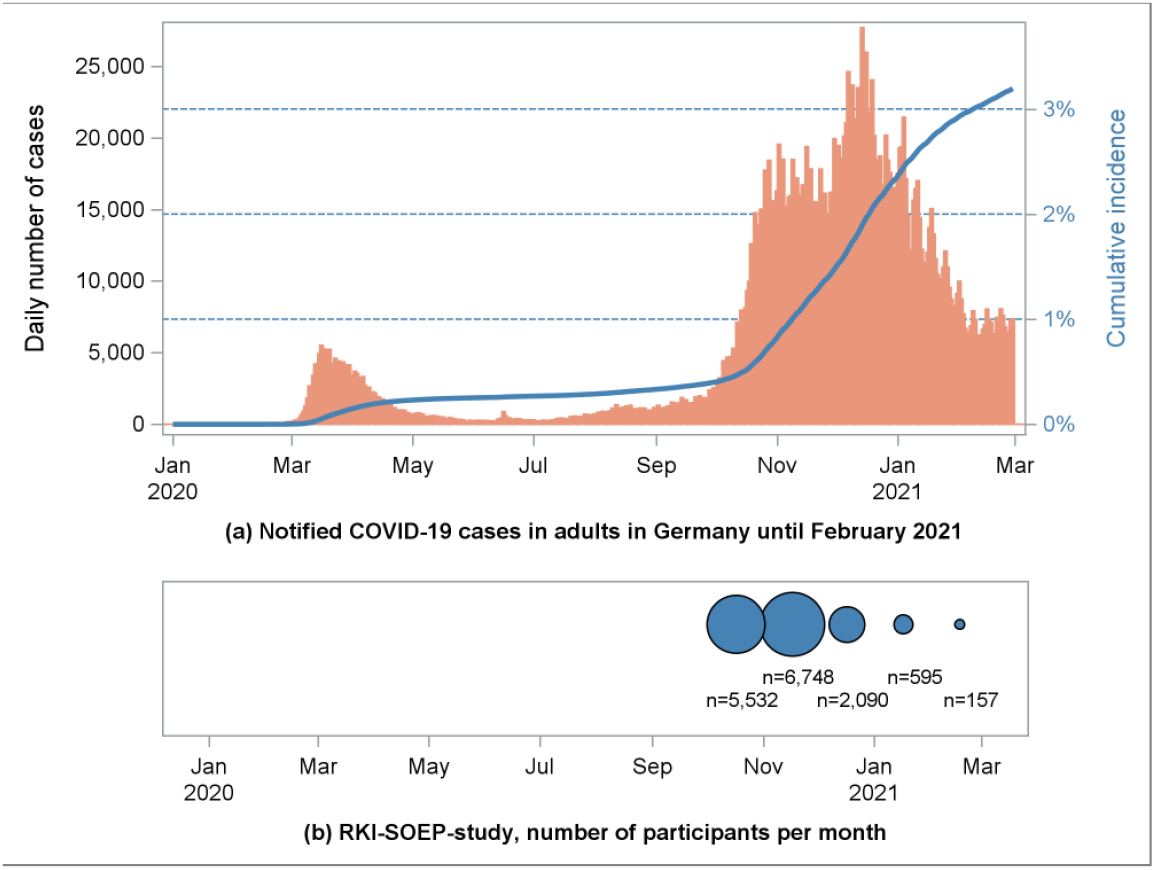
Notified COVID-19 cases in adults 18 years or older in Germany and sampling distribution of the RKI-SOEP study.

According to the nationwide sampling design, 31,675 adults from 19,569 households were invited to participate in the study and 15,122 adults from 9,781 households participated as shown in **Figure 2** (response 48%, American Association for Public Opinion Research response rate 6^15^). DBS specimens yielding valid laboratory test results were available from 14,781 participants (97.7% of all participants) and ONS from 97.1% of participants. Questions on pre-study SARS-CoV-2 testing (PCR_pre-study_) were answered by 98.6% of participants. The study sample included 146 participants with a self-reported pre-study positive SARS-CoV-2 test.

**Figure 2.**
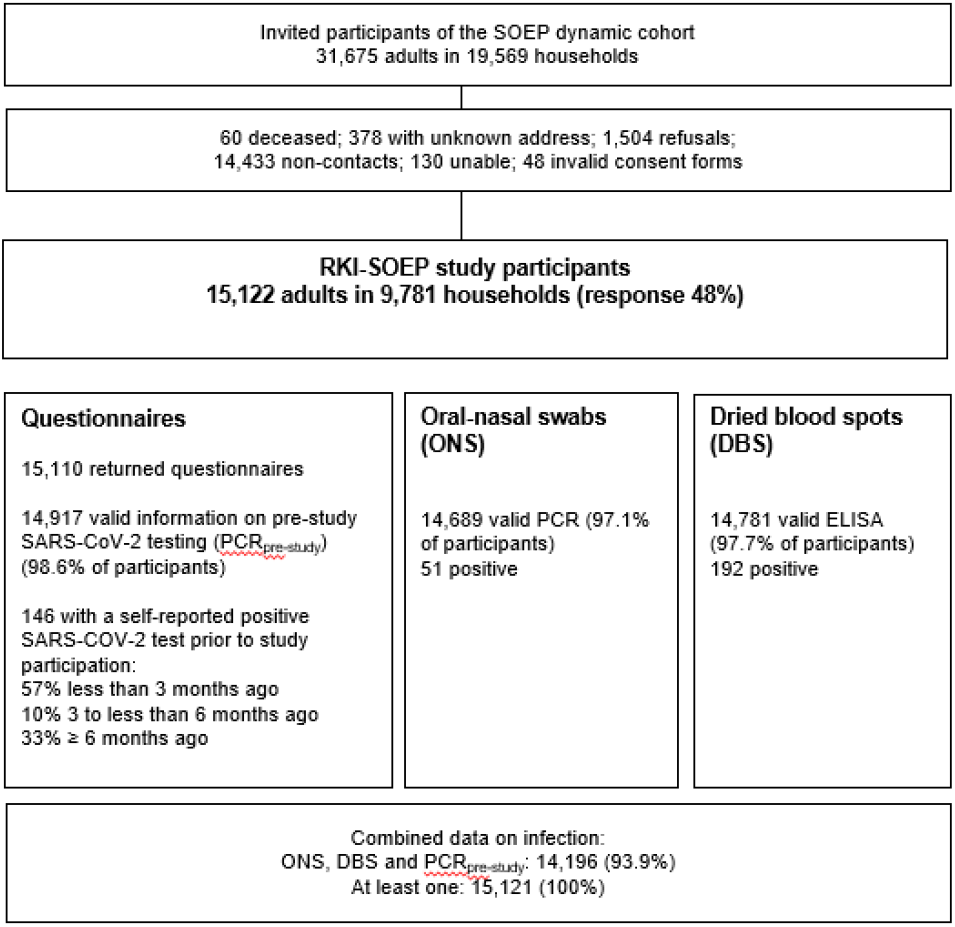
Flow-chart of study design of the RKI-SOEP study. Percent are unweighted.

We compared the observed and the expected proportion of self-reported laboratory-confirmed COVID-19 cases, and they were rather similar (**Figure 3**). The expected proportion was calculated from the cumulative incidence of notified non-deceased cases weighted by questionnaire completion date. The first positive PCR test occurred less than 3 months prior to study participation in 60% of cases, 3 to less than 6 months prior in 11%, and ≥ 6 months in 29% of cases (median time 70 days, interquartile range 33-193 days).

**Figure 3.**
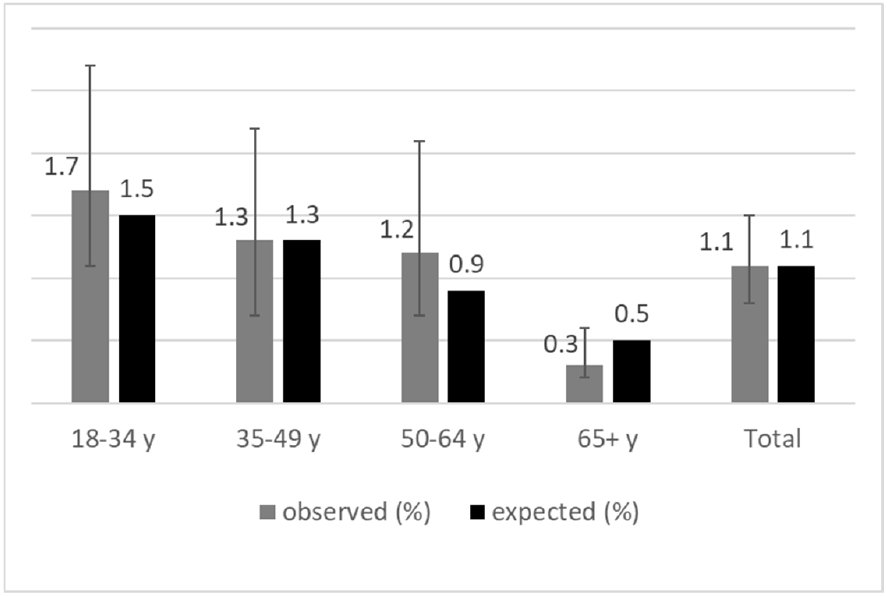
Observed (with 95% confidence intervals) versus expected proportion of notified COVID-19 cases in the study sample. Observed cases: self-reported prior positive SARS-CoV-2 tests. Expected cases: cumulative incidence of notified cases adjusted for questionnaire completion date and deceased cases.

The key socio-demographic characteristics of the study sample are shown in **Table 1** for all 15,122 participants and for subgroups of participants, i.e. for 146 participants with a self-reported positive SARS-CoV-2 test, 51 participants with a positive PCR test from study ONS (participants with an acute infection), 192 participants with a SARS-CoV-2-S1 IgG ELISA positive DBS test (seropositive participants), and 288 participants with past or current SARS-CoV-2 infections based on either one of the former infection categories. The 51 acute infections detected during the study corresponded to 0.4% of participants. Furthermore, **Table 1** shows the population-weighted proportion of participants with a prior or current SARS-CoV-2 infection which was 2.1% (95% CI 1.6-2.6%) for participants aged 18 to 99 years. In a multivariable logistic regression model, infection status was associated with larger households (> 4 persons), high socioeconomic deprivation at district level and higher incidence at district level (OR 2.03, 95% CI 1.03-4.00), but there was no statistically significant association with age group, sex or school education.

**Table 2** presents the prevalence of IgG antibodies against SARS-CoV-2. The 192 seropositive adults in the sample correspond to an unweighted seroprevalence of 1.3 % (95% CI 1.1 - 1.5%); 1.3 % (95% CI 1.0 - 1.7%) population-weighted; 1.3% (95% CI 0.9 - 1.7%) population-weighted and adjusted for test specificity of 0.997 and initial test sensitivity 0.811. Furthermore, study-specific sensitivity of 0.616 (95% CI 0.475 - 0.740) that takes into account antibody decay over time was estimated based on 133 participants with a self-reported positive SARS-CoV-2 test at least 11 days prior to DBS sampling. Population-weighted seroprevalence adjusted for specificity 0.997 and sensitivity 0.616 was 1.7% (95% CI 1.2 - 2.3%) and this is our main estimate of the cumulative seroprevalence in adults from the beginning of the pandemic in Germany to November 2020. Almost half (48%, 95% CI 37 - 59%) of seropositive participants were unaware of their infection.

We estimated a very similar proportion of undetected cases of 45% (95% CI 21 - 60%) in a separate analysis which compared the cumulative incidence of notified cases, sampling-density adjusted as described in the methods section, to the seroprevalence in our study, which was population-weighted and adjusted for test specificity 0.997 and for test sensitivity 0.616 that includes antibody decay (**Table 3**). This corresponds to an underascertainment ratio of 1.8 (95% CI 1.3 - 2.5). The point estimate for the underascertainment ratio was higher in districts with high socioeconomic deprivation (4.3, 95% CI 1.8 - 8.7) compared to mid-deprived (1.6, 95% CI 0.9 - 2.7) and low deprived (1.7, 95% CI 1.0 - 2.6) districts. Sensitivity analyses of the underascertainment ratio and of the proportion of undetected cases are shown in **Supplemental Table S3**, where we did not use study-specific information on antibody decay but incorporated findings from the literature on antibody decay in three scenarios: no decay; antibodies below detection threshold in one third of cases older than 4 months, and an extreme scenario assuming that all cases that occurred at least 6 months ago cannot be detected. Additionally, these sensitivity analyses restricted the age range to 18-69 in order to focus on the non-institutionalized population and also excluded fatal notified cases. Among these different scenarios, the underascertainment ratio ranged from 1.4 to 1.9 and the proportion of undetected cases from 26% to 47%.

A quarter of participants reported at least one SARS-CoV-2 test prior to the study (24%, 95% CI 22 – 25%) (**Table 4**). These tests were from nasal or oral swabs, the wording of the question excluded self-tests. They are assumed to be mostly PCR tests due to very limited availability of antigen tests during that time. Seropositive but infection-unaware participants reported a higher test frequency compared to seronegative participants (37%, 95% CI 22 - 54% vs. 23%, 95% CI 22 - 24%, p=0.06). They also reported to have been in contact with infected persons more often in comparison to seronegative participants. Generally, a higher proportion of women reported prior testing compared to men (25%, 95% CI 23 - 26% vs. 22%, 95% CI 21 - 24%; p=0.015). Women reported more often tests due to routine testing, e.g. occupational testing or routine testing on hospital admission, while men more often reported testing after travel return. The two oldest age groups had the lowest proportion of tested persons. Only 18% of adults with low school education had been tested compared to 25% and 26% of those with a medium and high level of school education, respectively (p<0.0001). Analogously, the test frequency decreased with higher district-level socioeconomic deprivation. Symptom-triggered test frequency was similar across educational groups. However, routine testing was more common in low-educated adults whereas travel-related testing and testing due to contact with an infected person was more common in highly educated groups. The test frequency in Bavaria was higher than in the other federal states of Germany, while in Brandenburg, Saxony and Saxony-Anhalt it was lower. The most frequent reason for previous testing was routine testing, e.g. occupational testing or aon hospital admission, and there were significant differences between the federal states.

**Table 4:** Self-reported frequency of SARS-CoV-2 tests since the beginning of the pandemic and reasons for testing (14,917 RKI-SOEP study participants with valid self-reports on pre-study tests, participation predominantly in October-November 2020)

|  |  | Proportion tested for SARS-CoV-2 prior to study |  | Reasons for testing*: Proportion of participants ever tested |  |  |  |  |  |  |  |
| --- | --- | --- | --- | --- | --- | --- | --- | --- | --- | --- | --- |
|  |  | Row % (95% CI), population-weighted |  |  |  |  |  |  |  |  |  |
|  | N | Ever tested | Tested more than once | Own symptoms | Contact to infected person | Contact to probably infected person | Routine test (e.g. occupational or pre-hospital admission) | On travel return | Prior to travel | Without special reason | Other reasons |
| Total | 14,909 | 24 (22-25) | 7.8 (7.1-8.6) | 27 (25-30) | 15 (13-17) | 8.3 (6.9-9.9) | 34 (32-37) | 21 (18-23) | 4.6 (3.4-6.1) | 8.7 (7.1-11) | 8.2 (6.8-9.9) |
| Serostatus |  |  |  |  |  |  |  |  |  |  |  |
| Seropositive and unaware of infection | 97 | 37 (22-54) | 20 (8.5-40) | 16 (5.2-39) | 35 (15-62) | 16 (5.4-40) | 15 (5.6-35) | 30 (8.9-65) | 13 (3.1-40) | 11 (2.7-36) | 4.7 (1.4-15) |
| Seropositive and aware of infection | 91 | 100 (-) | 47 (30-65) | 63 (46-77) | 49 (34-36) | 9.4 (4.1-20) | 13 (5.0-13) | 13 (5.3-27) | 1.5 (0.2-9.8) | 13 (5.0-30) | 4.0 (1.3-12) |
| Seronegative | 14,390 | 23 (22-24) | 7.4 (6.7-8.1) | 26 (24-29) | 13 (11-15) | 8.4 (7.0-10) | 35 (32-38) | 21 (18-23) | 4.6 (3.4-6.2) | 8.6 (7.0-10) | 8.3 (6.8-10) |
| Self-reported SARS-CoV-2 test |  |  |  |  |  |  |  |  |  |  |  |
| Tested positive | 145 | 100 (-) | 49 (34-64) | 54 (40-68) | 41 (29-55) | 10 (3.7-26) | 14 (7.3-26) | 17 (8.7-31) | 2.0 (0.5-7.8) | 13 (5.2-29) | 3.0 (1.2-7.4) |
| Tested negative or not tested | 14,764 | 23 (22-24) | 7.4 (6.7-8.1) | 26 (24-28) | 13 (11-15) | 8.2 (6.9-9.8) | 35 (33-38) | 21 (18-23) | 4.7 (3.5-6.3) | 8.5 (6.9-10) | 8.5 (7.0-10) |
| Sex |  |  |  |  |  |  |  |  |  |  |  |
| Women | 7,982 | 25 (23-26) | 8.8 (7.8-9.9) | 27 (24-31) | 15 (13-18) | 8.1 (6.4-10) | 38 (35-42) | 17 (14-20) | 3.6 (2.5-5.4) | 8.5 (6.7-11) | 8.5 (6.7-11) |
| Men | 6,927 | 22 (21-24) | 6.9 (5.9-7.9) | 27 (24-31) | 14 (11-17) | 8.6 (6.6-11) | 30 (26-34) | 25 (21-29) | 5.6 (3.7-8.4) | 8.9 (6.7-12) | 7.9 (5.9-10) |
| Age group in years |  |  |  |  |  |  |  |  |  |  |  |
| 18-34 | 2,778 | 29 (26-31) | 9.7 (8.1-12) | 31 (27-36) | 21 (17-26) | 8.3 (5.9-12) | 27 (22-32) | 24 (20-29) | 5.1 (3.0-8.6) | 9.3 (6.7-13) | 4.1 (2.4-6.8) |
| 35-49 | 3,523 | 31 (28-33) | 11 (9.3-13) | 35 (30-39) | 13 (10-16) | 11 (8.1-14) | 30 (26-35) | 21 (17-26) | 4.8 (3.0-7.6) | 8.6 (5.9-12) | 9.9 (7.2-13) |
| 50-64 | 4,885 | 22 (20-24) | 6.9 (5.7-8.4) | 22 (18-26) | 14 (11-19) | 8.0 (5.5-11) | 39 (34-44) | 19 (15-24) | 4.3 (2.5-7.3) | 7.9 (5.4-11) | 9.5 (6.9-13) |
| 65-79 | 3,057 | 15 (13-17) | 4.2 (3.3-5.5) | 11 (7.5-16) | 4.7 (2.6-8.5) | 4.2 (2.4-7.1) | 52 (44-59) | 14 (8.7-22) | 3.4 (1.6-7.0) | 9.4 (5.9-15) | 12 (7.7-17) |
| 80+ | 666 | 12 (8.7-16) | 3.2 (1.8-5.7) | 19 (6.0-45) | 1.0 (0.1- .1) | 2.8 (0.7-11) | 55 (36-72) | 15 (5.6-35) | 2.2 (0.4-12) | 7.2 (2.6-18) | 11 (4.6-22) |
| Household composition (< 60 y) |  |  |  |  |  |  |  |  |  |  |  |
| 1 person | 1,184 | 26 (22-29) | 9.1 (7.0-12.0) | 26 (20-34) | 17 (12-24) | 11 (6.7-17) | 31 (24-38) | 21 (15-28) | 9.2 (5.1-16) | 9.2 (5.7-14) | 8.2 (4.8-13) |
| 2-4 persons, incl. children ≤ 16 y | 3,538 | 30 (27-33) | 10 (8.5-12) | 35 (30-39) | 17 (14-22) | 11 (8.0-14) | 31 (27-36) | 19 (15-24) | 3.1 (1.7-5.4) | 9.6 (6.7-14) | 6.6 (4.5-9.4) |
| 2-4 persons, no children ≤ 16 y | 3,436 | 26 (24-29) | 8.3 (6.8-10) | 27 (22-31) | 15 (12-20) | 7.7 (5.2-11) | 31 (26-36) | 27 (21-33) | 4.2 (2.6-6.6) | 8.1 (5.4-12) | 6.3 (3.9-10) |
| > 4 persons, incl. children ≤ 16 y | 1,177 | 29 (24-36) | 11 (7.5-16) | 37 (27-48) | 21 (14-31) | 7.5 (4.2-13) | 23 (15-34) | 17 (10-26) | 6.0 (2.4-14) | 6.6 (3.3-13) | 11 (5.3-21) |
| > 4 persons, no children ≤ 16 y | 113 | 12 (5.9-22) | 4.9 (1.8-12) | 56 (29-80) | 20 (3.6-63) | 11 (2.4-40) | 12 (3.5-33) | 12 (3.7-34) | 0% | 0% | 0% |
| Not classified (60+ years) | 5,113 |  |  |  |  |  |  |  |  |  |  |
| **School education** |  |  |  |  |  |  |  |  |  |  |  |
| Low | 2,683 | 18 (16-20) | 5.7 (4.5-7.2) | 29 (23-36) | 9.9 (6.2-15) | 9.1 (5.8-14) | 46 (40-53) | 12 (7.9-18) | 2.4 (0.9-6.0) | 6.8 (4.2-11) | 10 (7.3-15) |
| Middle | 5,101 | 25 (23-27) | 8.0 (6.9-9.3) | 24 (21-28) | 12 (9.6-15) | 8.2 (6.1-11) | 34 (30-38) | 23 (19-28) | 3.7 (2.3-5.9) | 9.8 (7.1-13) | 9.2 (6.7-13) |
| High | 6,233 | 26 (24-28) | 8.5 (7.4-9.9) | 28 (25-32) | 17 (14-21) | 7.6 (5.8-9.8) | 32 (28-35) | 21 (17-24) | 7.2 (5.0-10) | 9.0 (7.0-12) | 6.6 (4.9-8.9) |
| **District incidence stratum\*** |  |  |  |  |  |  |  |  |  |  |  |
| High incidence (Cluster A) | 2,978 | 31 (28-34) | 12 (9.9-14) | 25 (21-31) | 16 (12-21) | 9.4 (6.6-13) | 30 (25-35) | 19 (15-24) | 8.8 (5.6-14) | 11 (7.4-16) | 11 (7.7-16) |
| Average incidence (Cluster B) | 8,572 | 23 (22-25) | 7.4 (6.5-8.4) | 28 (25-31) | 15 (13-18) | 7.5 (5.9-9.5) | 34 (31-38) | 22 (19-26) | 3.1 (2.1-4.6) | 8.4 (6.6-11) | 7.7 (6.1-9.7) |
| Late second wave (Cluster C) | 1,689 | 20 (17-24) | 6.6 (4.7-9.1) | 32 (25-40) | 9.8 (6.3-15) | 8.7 (5.1-14) | 43 (35-52) | 17 (11-25) | 2.3 (0.9-5.7) | 7.2 (4.0-13) | 5.3 (2.6-10) |
| Low incidence (Cluster D) | 1,665 | 16 (13-18) | 3.4 (2.5-4.7) | 22 (16-30) | 7.6 (4.1-14) | 10 (5.5-18) | 42 (33-51) | 17 (9.7-27) | 2.9 (1.0-8.4) | 4.2 (1.9-9.2) | 5.3 (2.7-10) |
| **Socioeconomic deprivation (district level)** |  |  |  |  |  |  |  |  |  |  |  |
| Low deprivation | 3,941 | 29 (27-32) | 9.7 (8.2-11) | 25 (21-30) | 16 (12-20) | 9.0 (6.6-12) | 32 (28-36) | 24 (19-29) | 5.9 (3.6-9.6) | 9.1 (6.2-13) | 10 (7.3-14) |
| Middle deprivation | 8,850 | 22 (21-24) | 7.4 (6.5-8.4) | 28 (25-32) | 14 (11-16) | 7.5 (5.9-9.4) | 35 (32-38) | 19 (16-23) | 4.0 (2.8-5.7) | 8.5 (6.6-11) | 7.5 (5.9-9.6) |
| High deprivation | 2,113 | 18 (16-21) | 5.8 (4.4-7.7) | 28 (21-35) | 14 (9.0-22) | 10 (6.5-16) | 42 (34-50) | 15 (10-23) | 2.6 (0.8-8.0) | 8.4 (4.9-14) | 5.5 (3.0-9.9) |
| **Federal state** |  |  |  |  |  |  |  |  |  |  |  |
| Schleswig-Holstein | 546 | 15 (11-20) | 3.9 (1.9-7.8) | 34 (20-51) | 7.6 (3.2-17) | 20 (9.4-37) | 27 (16-43) | 12 (5.4-24) | 4.4 (0.8-21) | 4.4 (1.2-15) | 13 (4.1-36) |
| Hamburg | 338 | 31 (23-39) | 6.7 (3.8-11) | 33 (22-47) | 15 (7.2-29) | 13 (5.3-28) | 26 (16-39) | 16 (6.4-36) | 6.9 (1.4-28) | 5.0 (1.9-13) | 9.0 (3.8-20) |
| Lower Saxony | 1,507 | 19 (15-22) | 4.7 (3.4-6.5) | 26 (19-34) | 14 (8.4-22) | 5.0 (2.6-9.1) | 36 (27-46) | 26 (17-38) | 2.1 (0.8-5.7) | 6.5 (3.4-12) | 4.0 (1.9-8.1) |
| Bremen | 119 | 23 (14-36) | 6.3 (2.7-14) | 39 (17-66) | 13 (4.1-33) | 5.9 (0.9-31) | 21 (7.2-46) | 26 (9.5-54) | 5.9 (0.8-32) | 1.5 (0.3-7.8) | 4.4 (1.2-15) |
| North Rhine-Westphalia | 3,080 | 26 (23-29) | 9.6 (8.0-12) | 27 (22-33) | 14 (10-19) | 5.8 (4.0-8.6) | 40 (34-46) | 15 (11-20) | 4.8 (2.7-8.3) | 6.3 (4.1-9.5) | 7.8 (5.2-11) |
| Hesse | 1,090 | 20 (16-25) | 4.3 (2.6-7.2) | 23 (15-34) | 12 (5.5-24) | 8.9 (4.5-17) | 23 (16-33) | 32 (21-46) | 2.8 (0.9-8.0) | 11 (4.5-26) | 11 (5.3-20) |
| Rhineland-Palatinate | 659 | 20 (15-25) | 6.5 (3.7-11) | 19 (11-30) | 18 (9.9-31) | 5.8 (2.4-13) | 42 (28-56) | 17 (9.0-30) | 1.8 (0.5-6.1) | 4.5 (2.0-9.5) | 6.9 (3.1-15) |
| Baden-Württemberg | 1,782 | 22 (19-25) | 7.0 (5.3-9.1) | 29 (22-36) | 17 (12-23) | 8.1 (4.8-13) | 31 (24-39) | 29 (21-37) | 4.2 (1.9-9.3) | 7.5 (4.4-13) | 4.1 (2.3-7.1) |
| Bavaria | 2,246 | 35 (32-39) | 14 (11-16) | 25 (20-30) | 15 (10-20) | 9.2 (6.2-14) | 33 (28-38) | 23 (17-29) | 5.0 (2.5-9.7) | 14 (10-20) | 12 (8.6-17) |
| Saarland | 131 | 21 (12-34) | 9.1 (3.4-22) | 21 (6.2-51) | 24 (7.0-56) | ( - ) | 69 (41-87) | 3.0 (0.7-12) | ( - ) | 0.1 (0.0-0.6) | 10 (2.7-31) |
| Berlin | 699 | 26 (21-32) | 11 (7.4-16) | 38 (26-52) | 20 (11-33) | 11 (5.0-21) | 23 (15-35) | 12 (6.9-21) | 12 (5.0-26) | 13 (6.4-26) | 9.0 (3.9-20) |
| Brandenburg | 556 | 15 (11-20) | 5.0 (3.0-8.3) | 27 (14-46) | 16 (7.0-32) | 13 (5.1-30) | 45 (29-62) | 6.8 (2.5-18) | 7.7 (1.4-33) | 3.7 (0.9-14) | 3.3 (1.3-8.0) |
| Mecklenburg-Western Pomerania | 310 | 18 (13-25) | 4.2 (1.9-8.7) | 25 (12-44) | 2.8 (1.0-7.9) | 6.3 (1.1-29) | 43 (25-63) | 20 (7.9-41) | ( - ) | 3.2 (0.7-14) | 10 (3.0-29) |
| Saxony | 946 | 18 (14-22) | 4.1 (2.6-6.6) | 38 (27-50) | 13 (7.4-20) | 10 (5.4-18) | 33 (23-45) | 19 (10-33) | 3.4 (1.3-8.6) | 1.2 (0.5-3.1) | 7.6 (3.7-15) |
| Saxony-Anhalt | 439 | 15 (11-21) | 3.7 (2.0-6.7) | 20 (11-34) | 14 (7.3-26) | 21 (8.7-44) | 33 (19-51) | 18 (8.7-35) | 5.6 (1.0-27) | 21 (9.6-40) | 3.1 (1.0-8.9) |
| Thuringia | 456 | 19 (13-27) | 6.5 (3.6-11) | 29 (16-47) | 6.3 (2.3-16) | 4.5 (1.5-13) | 55 (37-71) | 16 (6.3-36) | 1.3 (0.2-7.0) | 7.2 (1.7-25) | 3.2 (1.0-9.9) |

\*More than one answer possible.

## Discussion

This nationwide SARS-CoV-2 seroepidemiological study (RKI-SOEP study), among adults in private households in Germany, shows that by November 2020 only about two percent of adults in Germany had had a SARS-CoV-2 infection. This finding expands on an early analysis of the RKI-SOEP data which estimated a countrywide seroprevalence of 1.3% in adults but had not yet accounted for test characteristics and antibody decay over time ^16^. We estimate that in this first year of the pandemic, approximately a quarter of all adults had had at least one SARS-CoV-2 test and that slightly more than half of SARS-CoV-2 infections have been detected and notified. Corresponding with our finding that testing for SARS-CoV-2 was more common among advantaged socioeconomic groups, we found a higher rate of undetected cases among residents in socioeconomically deprived districts.

Our results on the low pre-vaccine SARS-CoV-2 seroprevalence in Germany are in line with other seroepidemiological studies with random samples from the general population in Germany – not including hotspot studies – up to November 2020, which also indicate relatively low seroprevalence rates^17^. Mostly, these were regional or local studies. The Rhineland study, testing from April to June 2020, found a seroprevalence of less than 1% in two districts of the city of Bonn ^18^, the STAAB-COVID program (June to mid-October) a seroprevalence of 1.3% in the city of Würzburg ^19^, and the KoCo-19 study representative of Munich showed 1.8% in the first (April to June 2020) and 3.3% in the second round (November to mid-December) ^20^. In the SaarCoPS study (until mid-October 2020), which is representative for the federal state of Saarland, the seroprevalence was around 1% ^21^. The MuSPAD study showed prevalences between 1.3% and 2.8% in different German regions between July and December 2020^22^. Of note, Munich, Saarland and some of the MuSPAD study locations tend to be more severely affected regions. There is only one Germany-wide study besides ours, Corona-BUND (August to mid-November 2020), with a seroprevalence of about 1% in the adult population. In its first round (July to August 2020), the study estimated that there were 1.8 times as many infections as reported by health authorities ^23^. While first local seroepidemiological studies in Germany, which were mainly conducted in hotspot areas, indicated underascertainment ratios of four to five ^2,7,24-26^, starting with the second half of 2020 underascertainment ratios were lower, in the majority of studies underascertainment was around two^22,23,27-31^. The Robert Koch Institute as the national Public Health Institute systematically tracks seroepidemiological studies in the general population as well as in special population groups conducted in Germany (www.rki.de/covid-19-serostudies-germany). Internationally, Germany can be classified among the countries with low seroprevalence in 2020, similar to Norway (0.9%, November to December 2020) ^32^, Denmark (2.0%, August to October)^33^, and, earlier, Iceland (0.3%, April to June) ^34^. Somewhat higher seroprevalences were found in nationwide population studies in Slovenia (4.3%, mid-October to mid-November 2020) ^35^, Spain (5.2%, April to June) ^36,37^, the Netherlands (4.5%, June to August) ^38^, and England (8.9%, November) ^39^ and a much higher seroprevalence of 28% in October-November was estimated for the Czech Republic^40^. In Norway, the estimated ratio between seroprevalence and cumulative incidence was 1.1 ^32^, the study from Iceland showed a ratio of 1.8 ^34^, and the Danish study a ratio of 6 in August and 3 in October and December ^41^. A recent systematic review of seroepidemiological studies worldwide with search between January and December 2020 showed an overall seroprevalence of 4.5%, the estimates being a median of 18.1 times higher than the corresponding SARS-CoV-2 cumulative incidence ^42^.

With regard to social disparities in infections with SARS-CoV-2, previous findings from national seroprevalence studies in the pre-vaccine era of the pandemic are inconsistent and sometimes contradictory^43-45^. A previous analysis of our RKI-SOEP data used information on vocational and academic qualifications in addition to school-leaving qualifications to assess the participants’ educational level, and found higher SARS-CoV-2 seropositivity in adults with lower education^16^. Taking this finding together with our present result that school education alone was not associated with seropositivity, it can be suggested that professional education is more crucial for the risk of infection than schooling. Occupational working conditions may be an important mediator in this relationship. For instance, lower-skilled workers in essential jobs may have had fewer opportunities to reduce occupational contact and mobility by working remotely during the pandemic than highly qualified academics.^46^ In this context, area-based patterns of infections need to be considered, as well. Previous ecological studies from Germany showed that notified SARS-CoV-2 infections shifted from more affluent districts at the very beginning of the pandemic to socioeconomically deprived districts in more advanced stages of the pandemic^47,48^. Especially in the second pandemic wave, accordingly, Germany’s most deprived districts had the highest rates of notified infections^47^.

The strengths of the RKI-SOEP study include the nationwide sampling covering 400 out of 401 districts in Germany, the embedding in a long-standing dynamic cohort with ample data that allows for sophisticated weighting and thus higher generalizability to the adult population, the user-friendly self-sampling methods accompanied by methodological studies for cutpoint adjustments and the perspective of a longitudinal follow-up. However, we could not include the institutionalized population and those with limited German language skills. Moreover, the possibilities for stratified analyses are limited due to the relatively small number of seropositive cases. Of note, the clustered structure of the sample (individuals within households) is considered by weighting both at household and individual level and by the analysis which takes the correlation within households into account. Therefore, the household-based sample does not introduce bias into the estimation of the seroprevalence and its variance.

It has been suggested that without a correction for the proportion of seroreversion, serological surveys underestimate the cumulative prevalence of infected persons in a population ^49^. The assessment of IgG antibody serum levels as a marker of a SARS-CoV-2 infection in serological surveys is limited by different long-term kinetics of SARS-CoV-2 antibodies depending on the target structure to which the antibodies are directed^50-52^, the applied laboratory assay ^51,53,54^, the severity of the disease^55,56^ and the time interval between exposure to the antigen (infection or vaccination) and blood sampling. Until now, long-term studies using the Euroimmun assay cover periods up to 9-months. Euroimmun assay-measured antibodies against the S1 subunit of the surface glycoprotein Spike-S show a decrease in antibody titers over time. Six to nine months after initial IgG seroconversion, results indicate a maintained seropositivity between 50% and over 80% ^57-60^. The maintained seropositivity of 62% after a median time of 70 days (interquartile range 33-193 days) in our study is lower which may be due to the population sample with more asymptomatic cases.

Testing has played a major role in Germany’s response to the pandemic ^61-63^. Regularly updated guidance on testing criteria has been issued^64^ (version history available at: https://edoc.rki.de/handle/176904/6459 and https://edoc.rki.de/handle/176904/6484.11). Rapid antigen tests started to become available towards the end of the fieldwork of our study. The social gradient in utilization of tests found in our study is in line with ecological evidence from Switzerland and from Massachusetts, where testing was associated with neighborhood socioeconomic position^65^ or neighborhood socioeconomic vulnerability index.^66^

Our findings of a lower test frequency and a higher underascertainment of cases in the socioeconomically most deprived districts of Germany suggest that testing-related disparities may have masked the magnitude of the social gradient in SARS-CoV-2 infections as found in previous ecological analyses of notification data.^47,48^ Although direct test costs to individuals were not involved in Germany, other barriers to testing access may exist, e.g. time and transport constraints, language or health literacy-related barriers. In addition, the distribution of the test frequency by federal state may reflect differences in the incidence, but also the different strategies of the federal states responsible for infectious disease control in Germany.

In conclusion, this study shows that shortly before the start of the German vaccination program only about 2% of adults in Germany had had contact with the virus and more than half of these cases had been detected and notified. This is a success for the containment and mitigation measures. At present (as of November 19^th^ 2021), Germany has 4.42 million notified cases in adults which corresponds to 6.4% of the population. Assuming an underascertainment ratio of around two among adults, as shown in this and other recent German studies, we estimate that almost 13% of the adult population in Germany has already had contact to the virus. Containment and mitigation measures as well as the vaccine campaign cannot reach their full potential for pandemic control if they are not accepted and followed by almost all of the population. However, only 68% of Germany’s population is fully vaccinated (78.6% among adults; November 21^th^ 2021), which, even considering an estimated underreporting of up to 5 percentage points^67^, is not sufficient and lags behind other Western European countries^68^. As the RKI-SOEP study set out to investigate social and regional aspects and factors that promote or hinder coronary infection and vaccine acceptance, it launched a second round in November 2021, again studying a representative sample of the population in Germany and this time differentiating vaccine- and infection-induced antibodies.

## Data Availability

The data cannot be made publicly available because informed consent from participants did not cover public deposition of data. However, the dataset underlying the analysis in this article is archived in the SOEP Research Data Centre (https://www.diw.de/en/diw_01.c.601584.en/data_access.html) in Berlin and can be accessed on site upon reasonable request.

## Acknowledgements

The authors commemorate in deep sorrow our recently deceased colleague Thomas Lampert who was one of the initiators of this study. The authors thank all persons who have contributed to the RKI-SOEP study, in particular Jörg Wernitz and Jennifer Allen (both RKI), Jan Goebel and Thomas Hollacher (both DIW/SOEP) and Parvati Trübswetter and Herbert Brücker (both IAB) for coordination and contributions to data security and legal issues; Markus Busch (RKI) for conceptual input to study design, ethical matters and questionnaire; Florian Griese (DIW/SOEP) for help with questionnaire layout; Tim Kuttig (RKI) for contributing to fieldwork concept and logistics; Rainer Siegers (DIW/SOEP) for assisting in sample definition and weighting; Martin Gerike, Marvin Petrenz, Jana Nebelin (all DIW/SOEP) and Andrea Männel (RKI) for data management; Jörg Schaarschmidt, Annett Klingner, Alexander Kroenke (all RKI) and Monika Wimmer (DIW/SOEP) for designing study information and documents; Silke Stahlberg, Antje Kneuer (both RKI) and the RKI laboratory teams involved in ONS and DBS analyses; Felicitas Vogelgesang, Stefan Junker and Stefan Damerow (all RKI) for statistical analyses presented in the Supplement; Heike Hölling, Isabell Hey, Gina Schöne and Jasmin Gundlach (all RKI) for quality assurance; Anja Engel (RKI) for assisting with table layout. We would also like to thank the entire team who established the SOEP study over several decades and the employees of Kantar GmbH who contributed to the field work and data collection of the RKI-SOEP study. We are grateful for the dedicated work of Heinrich Scheiblauer and his team at the Paul Ehrlich Institute, who conducted validation studies to establish the test performance. We sincerely thank all study participants for their willingness to participate.

## Author contributions

AG, LS, LW, TZ, JH, MMG, HB, SZ, SL made substantial contributions to the initiation, conception or design of the study. ASR conducted the analyses with substantial contributions from SZ and HWS. HN, MMG, SH, JH, JM, AN, CPM, FP, MS, HWS, HW, LS, AG contributed to the interpretation of data HN drafted the manuscript with substantial contributions from ASR, JH, FP, CPM, SZ, HWS. All authors critically revised the manuscript, edited and approved the final version.

The authors declare no competing interests.

