## Supplemental material for "Germany’s low SARS-CoV-2 seroprevalence confirms effective containment in 2020: Results of the nationwide RKI-SOEP study"

### Supplementary information

1. Application of the Euroimmun anti-SARS-CoV-2-S1-IgG ELISA antibody test to dried blood spots
2. Grouping of districts based on the incidence of notified SARS-CoV-2 infections over time
3. Methodological details and sensitivity analyses for the calculation of the underascertainment ratio and the estimated proportion of undetected cases.

### 1. Application of the Euroimmun anti-SARS-CoV-2-S1-IgG ELISA antibody test to dried blood spots

The qualitative Euroimmun Anti-SARS-CoV-2-ELISA (IgG) test for S1 antibodies has been most commonly used in the analysis of serum samples since early 2020. In the RKI-SOEP-study presented here, however, it is used to analyse dried blood spots (DBS). A method study was therefore conducted comparing serum with dried blood, embedded in the 'CORONA-MONITORING lokal' study [69]. This method study comprised 276 individuals who had participated both in the baseline survey in May/June 2020 and in the follow-up survey of the study 'CORONA-MONITORING lokal' in October 2020. The sample was made up of individuals who either had a positive or indeterminate IgG test result in serum measurements at the time of the baseline survey (n = 265) or had a negative test result but reported a positive PCR test before the baseline survey in the questionnaire (n = 11).

#### *Study execution and laboratory methods*

During the follow-up, the study team collected both a venous blood sample, which was processed into serum, and a capillary blood sample, which was processed into dried blood. Both samples were tested for IgG antibodies using Anti-SARS-CoV-2-ELISA (IgG) (Euroimmun AG, Lübeck, Germany, lot E200518BC). The results of this test are semiquantitative ratio values which were classified for serum samples using the manufacturer-supplied cutpoints (positive: ratio ≥ 1.1; indeterminate: 0.8 ≤ ratio < 1.1, negative: ratio < 0.8).

#### *Statistical analysis*

The aim of the analysis was to examine the test characteristics of the IgG test based on DBS compared to serum samples and, if appropriate, to derive a cutpoint adapted to dried blood so that the seroprevalence based on dried blood is comparable to a seroprevalence based on serum samples. The categorization used was 'positive' versus 'non-positive' (negative or indeterminate). Results of the serum measurement using the manufacturer-supplied cutpoints were regarded as the gold standard for the present analysis.

On the one hand, the adapted cutpoint was determined by minimizing the misclassification rate. To this purpose, cutpoints in the range 0.7−1.1 were used to classify the dried blood ratio values. This range was chosen since first analyses showed that dried blood spot samples yielded somewhat lower ratio values than serum samples. For each cutpoint, the proportion of misclassified DBS test results in comparison to serum results was determined, i.e. the proportion of all dried blood samples that were classified differently from the corresponding serum sample. Confidence intervals for the proportion of misclassified DBS test results were calculated using the Wilson score method [2,3].

On the other hand, a correction formula was estimated to predict serum ratio values from DBS ratio values, and the cutpoint was converted using this formula. The correction formula was estimated via piecewise linear regression, with the ranges for the piecewise regression defined by examining residual plots.

### *Results*

The measurements performed with dried blood (mean value 1.52, range 0.09 - 6.97) yielded slightly lower ratio values compared to the results from serum (mean value 1.68, range 0.11 - 6.72). Half of the serum samples collected in the follow-up survey were IgG positive (Supplemental Table S1).[1] Overall, the proportion of DBS samples misclassified was 5.1% compared to the corresponding serum sample, applying the manufacturer-supplied cutpoint to the DBS samples (14 of 276 dried blood samples were misclassified, 95% CI  3.0 - 8.3%) (see Supplemental Table S1). All misclassifications were false negative categorizations (10.1% of 138 positives in serum were categorized as negative in the DBS sample, 95% CI 6.1 - 16.3%).

***Supplemental Table S1:*** *Categorized IgG measurement in serum vs. categorized IgG measurement in dried blood spot using the manufacturer-supplied cutpoint (number, row percentage)*

| Result of serum sample | Result of dried blood spot sample | | |
|---|---|---|---|
| | Positive (≥ 1.1) | Non-positive (< 1.1) | **Total** |
| Positive (≥ 1.1) | 124 (89.9%) | 14 (10.1%) | 138 |
| Non-positive (< 1.1) | 0 (0%) | 138 (100%) | 138 |
| **Total** | 124 | 152 | 276 |

The minimum misclassification over all cutpoints tested was 2.9% (8 of 276 samples misclassified, 95% CI 1.5 - 5.6%). It was reached with a cutpoint of 0.94 and 0.95, respectively (see Supplemental Table S2; the categorizations for these two cutpoints were identical). With this cutpoint, false positive and false negative misclassifications occurred with equal frequency.

***Supplemental Table S2:*** *Categorized IgG measurement in serum vs. categorized IgG measurement in dried blood spot using the cutpoint that minimizes the overall misclassification rate (number, row percentage)*

[1] Differences to the baseline IgG categorization may be explained by two factors: (1) waning of antibodies between baseline and follow-up; (2) use of a different test batch.

| Result of serum sample | Result of dried blood spot sample | | |
|---|---|---|---|
| | Positive (≥ 0.94) | Non-positive (< 0.94) | **Total** |
| Positive (≥ 1.1) | 134 (97.1%) | 4 (2.9%) | 138 |
| Non-positive (< 1.1) | 4 (2.9%) | 134 (97.1%) | 138 |
| **Total** | 138 | 138 | 276 |

As another way to establish a cutpoint, a correction formula was derived to convert the DBS values into serum values. This resulted in a good model fit when using piecewise linear regression:

(1) For DBS values < 0.19 (n = 8):

predicted serum ratio value = DBS value

(2) For DBS values from 0.19 to 2.2 (relevant range for the categorization into positive/negative), the following applies (n=201):

predicted serum ratio value = 0.074 + 1.093 × DBS ratio value

The explained variance ($R^2$) in this range is 95.5%. The intercept (0.074) has a standard error of 0.0169, and the slope parameter (1.093) has a standard error of 0.017.

(3) For DBS values > 2.2 the following applies (n=67):

predicted serum ratio value = 0.166 + 1.013 × DBS ratio value

The explained variance ($R^2$) for these high DBS values is 92.1%. The intercept (0.166) has a standard error of 0.015 and is therefore not significantly different from zero. The slope parameter (1.013) has a standard error of 0.037 and is not significantly different from 1.

Supplemental Figure S1 (left panel) shows the data points together with the estimated regression line. The right panel of the figure examines the agreement between measured serum ratio values and the serum ratio values predicted from the regression on DBS ratio values, using a Bland-Altman plot [4]. For this plot, the difference between the measured value and the predicted value is plotted against the mean of the two values. The plot indicates a uniform distribution of differences around zero throughout the range of values with only a small number of outliers, indicating a good model fit.

According to the correction formula (2), a DBS ratio value of 0.94 corresponds to a serum ratio value of 1.1 (by inverting the above regression equation: (1.1 − 0.074)/1.093 = 0.939). Thus, this method yields an adjusted cutpoint of 0.94 for dried blood spot samples.

### *Implementation in the analysis of the seroprevalence study*

Both using the correction formula and by minimizing the misclassification rate, 0.94 is obtained as the adapted cutpoint for classifying dried blood spot samples as IgG positive. This cutpoint was therefore used in the evaluation of the RKI-SOEP seroprevalence study to classify the ratio values of the Euroimmun IgG antibody test in dried blood spot samples.

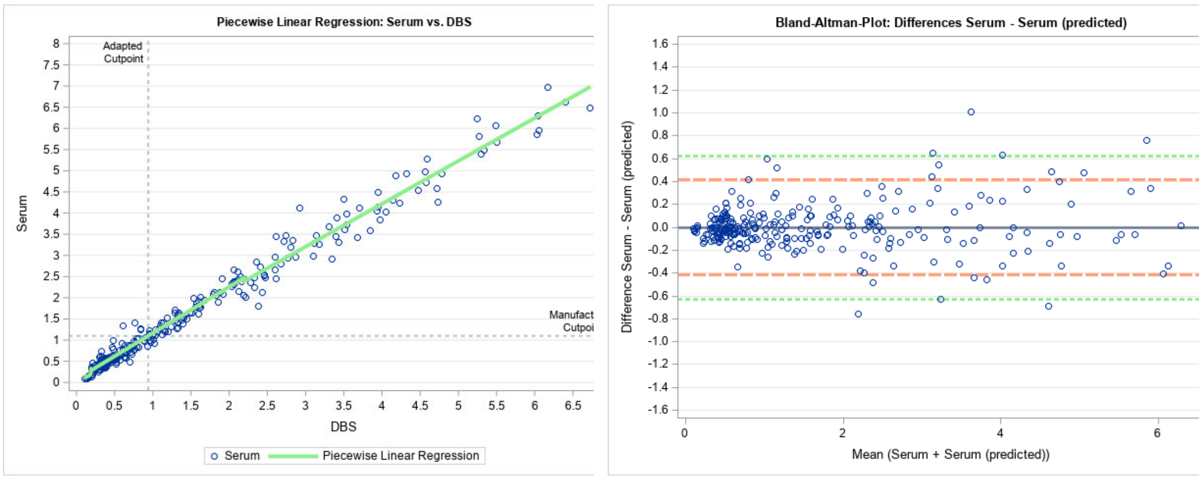


***Supplemental Figure S1: Left panel:*** *Data points and piecewise regression line for the regression of serum IgG ratio values on DBS IgG ratio values.* ***Right panel:*** *Bland-Altman plot of the difference between the measured serum IgG ratio value and the value predicted by the regression model against the mean of the two values. The lines show the limits of agreement (red, dashed line: ± 2 standard deviations; green, dotted line: ± 3 standard deviations).*

#### *References*


[1] Santos-Hövener C, Busch MA, Koschollek C et al. (2020) Seroepidemiological study on the spread of SARS-CoV-2 in populations in especially affected areas in Germany – Study protocol of the CORONA-MONITORING lokal study. Journal of Health Monitoring (S5/20): 1-18

[2] Wilson EB (1927) Probable inference, the law of succession, and statistical inference. Journal of the American Statistical Association 22: 209-212

[3] Newcombe RG (1998) Two-sided confidence intervals for the single proportion: Comparison of seven methods. Statistics in Medicine 17: 857-872

[4] Bland JM, Altman DG (1999) Measuring agreement in method comparison studies. Statistical Methods in Medical Research 8: 135-160


### 2. Grouping of districts based on the incidence of notified SARS-CoV-2 infections over time

The aim of the analysis described here was to derive a regional stratification within Germany, forming strata of districts with a similar distribution of notified COVID-19 cases over time.

#### *Data used*

The data set comprises the weekly 7-day incidence of notified laboratory-confirmed SARS-CoV-2 infections per 100,000 inhabitants for each of the 401 German districts. Data was extracted on 2021-03-21 from the internal server of the Robert Koch Institute, which hosts the mandatory infectious *disease* notifications in Germany. The data set comprises the incidence from calendar week 5 in 2020 (end of January) when the first COVID-19 case in Germany was reported to calendar week 6 in 2021 (mid-February). This end date was chosen as study participation continued until the end of February 2021 and we assume IgG antibodies can be detected on average 14 days after symptom onset.

### *Statistical analysis*

The 7-day incidence of SARS-CoV-2 infections was categorized from very high incidence (200 cases and more per 100,000) over high incidence (100 to less than 200 cases per 100,000), elevated incidence (50 to less than 100 cases per 100,000), medium incidence (15 to less than 50 cases per 100,000) to low (<15 per 100,000) and zero incidence (no new cases per 100,000).

In order to build trajectories over time, so-called (temporal) sequences were built for each of the 401 districts, displaying their time-ordered categorized weekly incidence in a sequence index plot. Figure S2 shows the temporal sequences for 10 districts as an example. Of note, the term "sequence" here refers to the time-ordering of the incidences, and its meaning is different from a genetic sequence in molecular epidemiology.

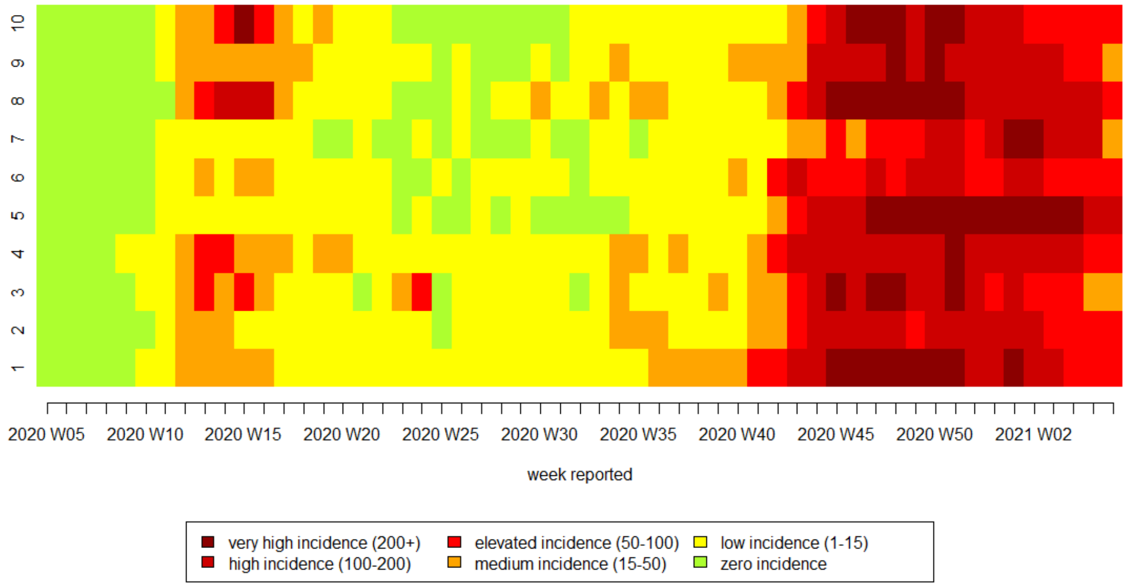


***Supplemental Figure S2:*** *Temporal sequence index plot: Weekly new COVID-19 cases per 100,000 for ten exemplary districts*

To form groups of districts with similar patterns over time, a so-called sequence analysis was performed using 'TraMineR 2.2-1' [1,2] in the R statistical package, version 4.0.5 (R Core Team 2021, R Foundation for Statistical Computing, Vienna, Austria). Pairwise distances between the temporal sequences were calculated using optimal matching. Using the distance matrix obtained from optimal matching, a statistical cluster analysis was performed using the 'WeightedCluster 1.4-1' [3] and the 'cluster 2.1.1' package [4]. Best clustering method and optimal number of clusters were evaluated using the 'wcCmpCluster' function. We chose 4-medoids clustering with the PAM algorithm, determining the start medoids with hierarchical Ward clustering. The statistical clusters found through this analysis are referred to as "district incidence strata" in the main text of the manuscript.

### Results

Plotting all temporal sequences (first unordered as a temporal sequence index plot (Supplemental Figure S3, left panel) and second ordered as temporal sequence density plot (Supplemental Figure S3, right panel)) shows that the data captures the complete first wave of the SARS-CoV-2 pandemic in Germany and almost the complete second wave.

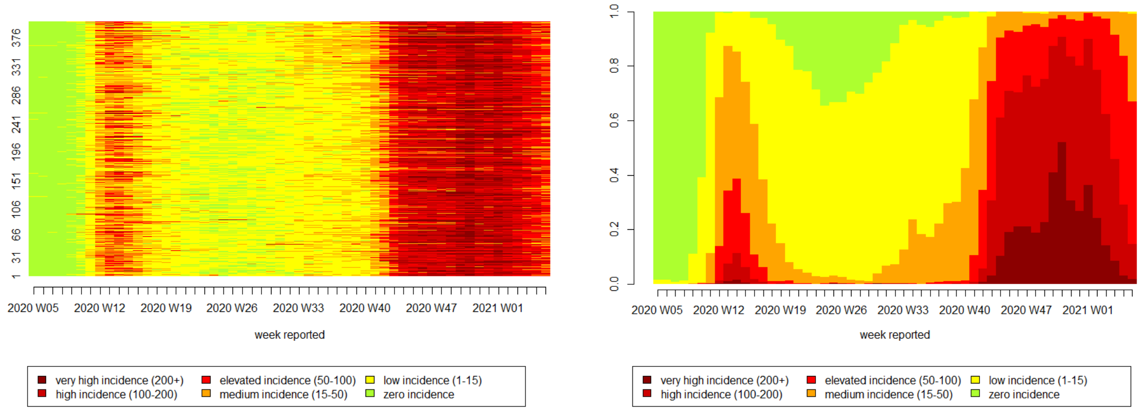


***Supplemental Figure S3: Left panel:*** *Temporal sequence index plot: Weekly new COVID-19 cases per 100,000 for each of the 401 districts in Germany, January 2020 to mid-February 2021.* ***Right panel:*** *Temporal sequence density plot: Weekly distribution of the COVID-19 incidence categories, January 2020 to mid-February 2021.*

Based on this data, four statistical clusters (groups of districts) were formed (Supplemental Figure S4). Cluster A ('high incidence') includes 50 districts. These districts were all at least affected with medium incidence in the first wave and were strongly and for a long time confronted with high and very high incidences during the second wave. Cluster B has a similar distribution in the first wave, but these 223 districts were less strongly affected in the second wave. As it captures the majority of the districts, it is called 'average incidence cluster'. The last two statistical clusters had lower incidences during the first wave. In the second wave, Cluster C (67 districts) was highly affected, similar to Cluster A, but with a later onset compared to the first two clusters. It is therefore called 'late second wave'. Cluster D (61 districts, 'low incidence') had lower incidences in both waves compared to the other clusters. Supplemental Figure S5 shows the regional distribution of the four statistical clusters in Germany.

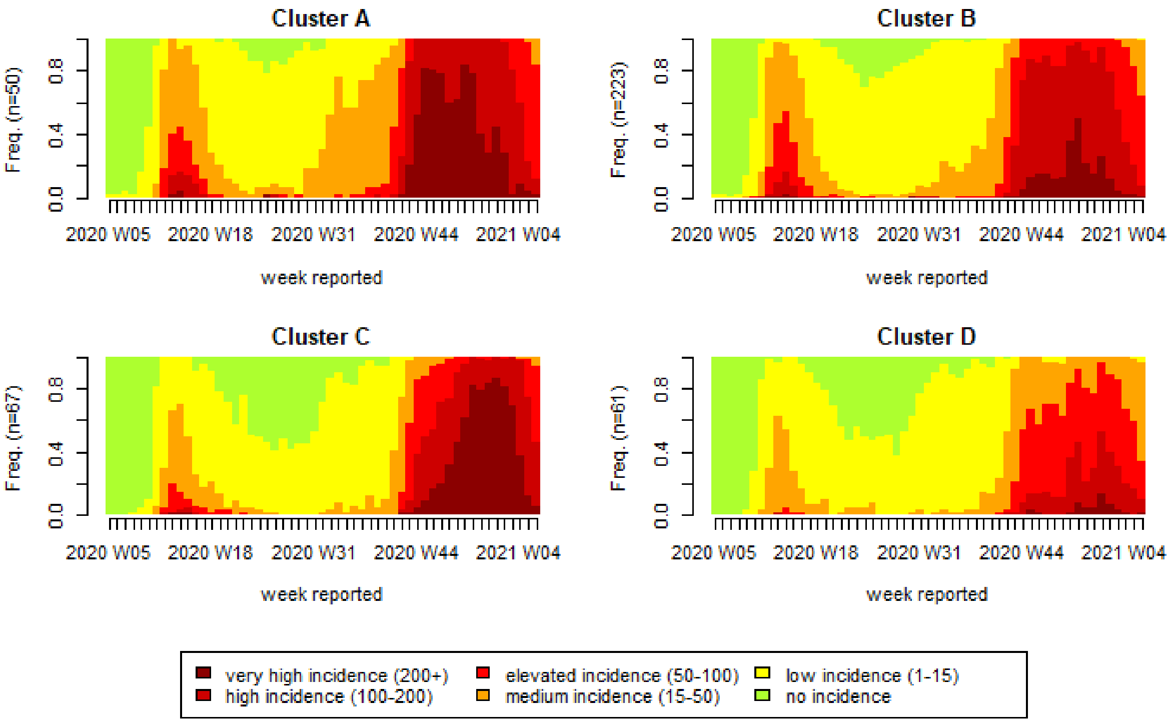


***Supplemental Figure S4:*** *Temporal sequence density plots for the four statistical clusters, 401 districts in Germany, January 2020 to mid-February 2021*

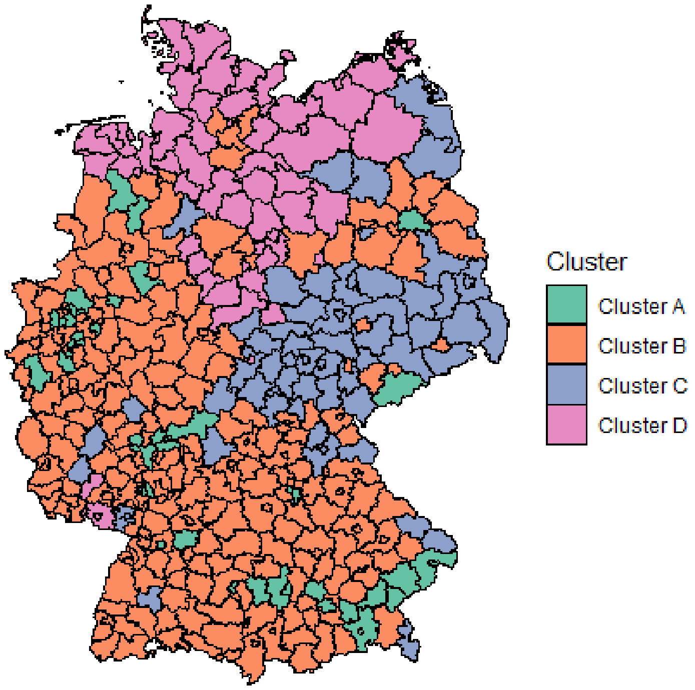


***Supplemental Figure S5:*** *Regional distribution of the four statistical clusters in Germany, January 2020 to mid-February 2021*

### *References*

[1] Abbott A, Tsay A (2000). “Sequence Analysis and Optimal Matching Methods in Sociology,Review and Prospect.”Sociological Methods and Research,29(1), 3–33

[2] Gabadinho, A., Ritschard, G., Müller, N. S., & Studer, M. (2011). Analyzing and Visualizing StateSequences in R with TraMineR. Journal of Statistical Software, 40(4), 1-37. DOI 10.18637/jss.v040.

[3] Studer, Matthias (2013). WeightedCluster Library Manual: A practical guide to creating typologies of trajectories in the social sciences with R. LIVES Working Papers, 24. DOI: 10.12682/lives.2296-1658.2013.24.

[4] Maechler M, Rousseeuw P, Struyf A, Hubert M, Hornik K (2021). cluster: Cluster Analysis Basics and Extensions. R package version 2.1.2.

### 3. Methodological details and sensitivity analyses for calculation of the underascertainment ratio and the estimated proportion of undetected cases.

The number of infections missed by the mandatory notification system was estimated in two ways: first internally, by estimating the proportion of seropositive cases that was unaware of the infection (main text **Table 2**); and second by comparing the observed seroprevalence, adjusted for test characteristics, to the number of notified cases, adjusted for sampling density (main text **Table 3** and **Supplemental Table S3**).

To this end, we calculated the cumulative incidence of notified cases individually for each participant, counting notified cases with symptom onset until DBS sampling date minus 14 days. If symptom onset was missing in notified cases, it was imputed by the median based on age, calendar week, federal state and day of the week (overall median: 4 days from symptom onset to notification date). In the main analysis, only cases with non-fatal disease course were included, as deceased cases had no opportunity to participate in the study. In sensitivity analyses (**Supplemental Table S3**), all cases were included. Next, the underascertainment ratio (ratio of seroprevalence to cumulative incidence of notified cases) and the estimated proportion of undetected cases were calculated (seroprevalence minus cumulative incidence of notified cases, divided by seroprevalence). In these calculations, we used the internally estimated sensitivity of 0.616, as this accounts for antibody decay over time as observed in the study population and thus represents the most appropriate assumption in our view. In addition, **Supplemental Table S3** shows results adjusting seroprevalence for the initial test characteristics, but with three different assumptions on antibody decay implemented in calculating the cumulative incidence of notified cases: (a) no antibody decay over time, (b) loss of seropositivity in 1/3 of infected persons 4 months after (reported or imputed) symptom onset, (c) loss of seropositivity in *all* infected persons 6 months after (reported or imputed) symptom onset.

**Supplemental Table S3:** Sensitivity analyses for the underascertainment ratio and the estimated proportion of undetected cases

| | | Sensitivity analyses | | | Base case |
|---|---|---|---|---|---|
| | | **Seroprevalence:** population-weighted and adjusted for initial test characteristics | | | **Seroprevalence:** population-weighted and adjusted for specificity = 0.997 and sensitivity = 0.616 that includes antibody decay observed in the study |
| | **Assumption on antibody decay over time:** | **Notified cases: (a) as is** | **Notified cases: (b) 1/3 of notified cases older than 4 months discounted** | **Notified cases: (c) all notified cases older than 6 months discounted** | **Notified cases: (a) as is** |
| **Population age group** | **Notified cases considered** | **Cumulative incidence of notified cases*** | | | |
| **18-99 yrs** | All cases | 0.9% | 0.8% | 0.7% | 0.9% |
| | Non-fatal cases | 0.9% | 0.8% | 0.7% | 0.9% |
| **18-69 yrs** | All cases | 1.0% | 0.9% | 0.8% | 1.0% |
| | Non-fatal cases | 1.0% | 0.9% | 0.8% | 1.0% |
| | | **Underascertainment ratio (95% CI)*,**** | | | |
| **18-99 yrs** | All cases | 1.35 (0.9 – 1.9) | 1.51 (1.1 – 2.1) | 1.82 (1.3 – 2.5) | 1.78 (1.3 – 2.5) |
| | Non-fatal cases | 1.38 (1.0 – 1.9) | 1.54 (1.1 – 2.1) | 1.85 (1.3 – 2.6) | 1.82 (1.3 – 2.5) |
| **18-69 yrs** | All cases | 1.43 (1.0 – 2.0) | 1.58 (1.1 – 2.2) | 1.88 (1.3 – 2.6) | 1.88 (1.3 – 2.6) |
| | Non-fatal cases | 1.44 (1.0 – 2.0) | 1.59 (1.1 – 2.2) | 1.88 (1.3 – 2.6) | 1.90 (1.3 – 2.7) |
| | | **Proportion of undetected cases (95% CI)*,***** | | | |
| **18-99 yrs** | All cases | 26% (5 – 47) | 36% (8 – 55) | 45% (22 – 60) | 44% (20 – 60) |
| | Non-fatal cases | 28% (3 – 48) | 35% (7 – 53) | 46% (23 – 61) | 45% (22 – 60) |
| **18-69 yrs** | All cases | 30% (1 – 50) | 37% (9 – 55) | 47% (23 – 62) | 47% (24 – 62) |
| | Non-fatal cases | 30% (0 – 50) | 37% (10 – 55) | 47% (23 – 62) | 47% (24 – 62) |

*Sampling density adjusted, i.e. each participant contributes according to the cumulative incidence of notified cases with symptom onset (notified or imputed) corresponding to his/her DBS testing date minus 14 days, discounting cases with symptom onset more than 4 months or more than 6 months before DBS testing as indicated.
**Underascertainment ratio: Ratio of seroprevalence to cumulative incidence of notified cases.
***Proportion of undetected cases: Seroprevalence minus cumulative incidence of notified cases, divided by seroprevalence.
****NA = estimate not available
